# Clinical Prediction Model Performance Metrics Cluster at Arbitrary Thresholds and are Incorrectly Reported in High-Ranking Journals: An Observational Meta-Research Study

**DOI:** 10.64898/2026.11.02.26367135

**Authors:** Alexander D Gibson, David N Borg, Nicole M White, Adrian G Barnett

## Abstract

**Background:** Previous research has shown that clinical prediction models have likely been “hacked” to meet arbitrary performance thresholds. However, this work only assessed abstracts, could not identify models with a perfect AUC (area under the receiver operating characteristic curve) of 1, and did not examine other performance metrics.

**Methods:** We collected data from full texts and validated a large language model to extract data from a random sample of Q1 journal articles for 2025. We examined if hacking occurred in the AUC value, sensitivity and specificity, and checked if articles correctly reported the AUC value from corresponding receiver operating characteristic (ROC) curves.

**Results:** We identified 2,001 prediction model articles that reported 15,656 AUC values. We found a striking excess of results near perfect prediction and at multiples of five and ten for the AUC, sensitivity and specificity, indicating likely manipulated data, methods or publication bias. Around 1 in 40 AUC values were reported outside of their confidence intervals, and around 1 in 8 ROC curves did not match their reported AUC value.

**Conclusion:** We have shown that not only the AUC value, but the sensitivity and specificity are being hacked, likely to help the authors publish their paper at the cost of evidence for practice. Excess models that are not representative of their true performance may misguide clinicians, distort the literature, and make it harder to find prediction models that are useful for practice.

## 1.0 Introduction

Clinical prediction is a rapidly growing field of research, with nearly 250,000 new models published between 1950 and 2024 (1). Extensive problems exist in current publications, including poor reporting, and inappropriate data or methods leading to many wasteful studies (2–8). As stark examples of poor practice, a statistician-led review found only 7 of 606 published COVID-19 prognostic models were of sufficient quality for clinical practice, while a sepsis model used in hundreds of United States Hospitals missed two-thirds of patients with sepsis (9,10). These issues contribute to research waste and misguided predictions which can lead patients to receiving incorrect treatments or not receiving necessary treatments.

The area under the receiver operating characteristic curve (AUC) value, is a widely used metric of a model’s ability to discriminate between events (e.g., death versus survival). The AUC value ranges from 0.5 (a coin toss) to 1.0 (perfect discrimination). Arbitrary thresholds have been purported at 0.7, 0.8 and 0.9, providing incentives for researchers to target these thresholds (4,11). This incentive may tempt researchers to potentially ‘hack’ their data, methods or reporting to inflate these values for their models (12). The appropriate design and reporting of models is necessary for transparency and implementation into practice, with various expert-led guidelines being published (13–15). Unfortunately, there is little evidence these guidelines have been widely implemented (16).

Recently, researchers examined the distributions of AUC values, finding evidence of potentially inflated model performances (4). They were shown to have excess results around arbitrary thresholds, indicating potential publication bias and researchers inflating model performances— termed AUC-hacking (4). However, the study only assessed values reported in abstracts where researchers may report the best performing models, and the study could not assess models with an AUC of 1.0. Additionally, no other performance metrics were examined, nor was it checked if the AUC values were correctly reported against the receiver operating characteristic (ROC) curve

Publish or perish culture has spurred questionable research practices in psychology, ecological and evolutionary research, and biomedical research (17–19). If these practices are also widespread in prediction model research published in high ranking journals it is likely to be consequential to outcomes as these journals generally receive more readers and are therefore more likely to influence practice (20,21). As previous research has shown, model development studies produce larger AUC values than those in subsequent validation studies (22–26), indicating there may be substantial and clinically meaningful differences between a model’s numeric and clinical performance.

This study aimed to examine if hacking practices exist in recently published clinical prediction models from Q1 journals. Our primary aim was to check for AUC-hacking in recently published journal article full texts. We also examined if reported AUC values corresponded with AUC values of the ROC curve, and if there is evidence that sensitivity and specificity values are also hacked.

## 2.0 Methods

We assembled a representative sample of recently published clinical prediction model articles. Journal rankings were downloaded from *Scimago* (https://www.scimagojr.com/journalrank.php) for 2024 on January 13, 2026. Results were filtered to Q1 ranked journals. The *PubMed* ID, title and abstract text were extracted from *PubMed* using the *rentrez* package (27) in *R* (version 4.6.1). The search date was January 13, 2026.

The search returned 63,954 articles. Our protocol and analysis plan were registered on Feb 5, 2026 (28). This study didn’t require ethics approval as it used publicly available data and there are no risks to data collection.

### 2.1 Search Filter

We used a custom filter to gather a representative sample of discrimination performance values from published prediction models. The AUC value and its commonly substituted terms were searched in the title and abstracts of published articles in *PubMed* (Supplementary 1).

### 2.2 Inclusion and exclusion criteria

Articles were included if they were, published in a quartile one journal, had at least one clinical prediction model, and were published in 2025. Articles were excluded if they were: pharmacokinetic studies which frequently mention “Area under curve” (AUC), meta-analyses, systematic reviews, pooled analyses or tutorial articles; the full text was not accessible; the article was not in English; or was an animal study.

### 2.3 Screening for study inclusion

Article screening for eligibility was completed independently, in duplicate, by AG and DB using the software *Rayyan* (29). Agreement was measured using Gwet’s AC1 agreement statistic (30). First, 10% of the articles were manually screened by both authors. Discrepancies were discussed between the two reviewers for inclusion or exclusion. Gwet’s AC1 statistic was > 0.8, so, as defined in our protocol (31), AG screened the remaining articles after discussions, to reduce overall screening time. (31).

### 2.4 Data Extraction

Data from 176 articles were extracted manually and compared to an extraction by a large language model (LLM; Claude Sonnet 4.6) (see Supplementary 2 for LLM prompt). LLMs have previously shown accurate performance for extracting data from published papers (32). Both the LLM and manual extractions collected the same set of information from the full texts.

Data extracted from full texts were: the AUC, sensitivity and specificity values including confidence intervals, sample size, and number of AUC values. We also recorded if the model was diagnostic or prognostic, using a developmental or validation study, and if there was an ROC curve.

We validated the LLM by manually extracting data from 176 (9%) articles. We checked accuracy in extraction by examining 50 values that differed between the manual and the LLM extraction.

These included 50 values where the manual extraction retrieved a value, but the LLM didn’t, and 50 values where the LLM extraction retrieved a value, but manual extraction didn’t. We primarily examined if any data were incorrectly extracted.

### 2.5 Statistical methods

Reported AUC values were visualised using a histogram with bin widths of 0.01 on a scale of 0.5 to 1.0 while sensitivity and specificity were visualised with bin widths 1 percent on a scale from 0% to 100%. For example, a bin width of (0.78, 0.79] included values > 0.78 and values ≤ 0.79. Three smooth estimates of the underlying distribution were fit for the AUC, using a b-spline basis with two to four degrees of freedom. Each of the models were fit with a random intercept to adjust for repeated values from the same article and used a negative binomial distribution. (4,33). The best fitting degrees of freedom was assessed by the Akaike information criterion (AIC) and was used to calculate the residuals. These residuals were visualised in separate histograms. Alternative models varying the b-spline boundary knots beyond the AUC value limits were used to reduce any fitting artifacts at an AUC of 1 (4,33).

### 2.7 ROC curves

Data from available ROC curves were extracted via a semi-automated process using the *PlotDigitizer* software (version 3.3.9 Pro). The *PlotDigitizer* software has been previously validated as a reliable tool for data extraction (34). We cross-checked the reported in-text AUC value against the AUC value extracted from the ROC curve. The AUC value was calculated using a trapezoidal function from the extracted coordinates of the ROC curve in the R package *bayestest* (35).

Differences between reported AUC values and their corresponding value extracted from ROC curves were plotted using a histogram. Bin widths for the histogram were 0.005. We examined the number of articles with differences between the reported and extracted AUC value of ≥ 0.02 which would provide evidence of altered reporting. A symmetric distribution centred on zero would indicate no overall bias. A skewed positive bimodal distribution would indicate researchers are altering the AUC value in their reporting and was our primary indicator of AUC-hacking by reporting.

All analyses were undertaken in *R* (version 4.6.1). All data and code are publicly available on *GitHub* (https://github.com/alexdgibson/cpm_auc) (36).

## 3.0 Results

Screening agreement between the two reviewers for the first 220 articles was 0.83 (95% CI = 0.68 to 0.84). The remaining articles were screened by one reviewer, as this met our a priori agreement criteria above 0.8 (28). There were 672 additional articles included (34%) that met the inclusion criteria that were inadvertently missed during initial filtering for journal articles. These articles were not examined in the ROC curve extractions or validation checks but were reported for all other statistics.

After screening, there were 2,166 out of 3,996 articles remaining. One-hundred and seventy articles were excluded during full text data collection because they did not contain the required information or were not clinical prediction modelling studies. There were 2,001 articles included for analysis. From these 2,001 articles, there was a total of 15,656 AUC values extracted, 7,600 sensitivity values from 1,213 of the 2,001 (61%) articles, and 7,176 specificity values from 1,161 of the 2,001 (58%) articles.

Confidence intervals were reported for 9,002 of the 15,656 AUC values (58%) in 1,329 of the 2,001 articles (66%). There were 227 AUC values that were outside the associated confidence interval (2.5%) or around 1 in 40. The mean (SD) sensitivity value was 74 (SD = 19) and the mean specificity value was 77 (SD = 17). The median number of AUC values per article was 5 (1^st^ to 3^rd^ quartile = 3 to 10). No AUC values below 0.0 were reported, but there was one AUC value erroneously reported above 1.0.

There were large excess results at (0.99,1.0] for the AUC value and (99,100] for sensitivity and specificity values (Figure 2, 3 and 4). A large underrepresentation of values at (0.90, 0.99] were seen for the AUC value and (90, 100] for both the sensitivity and specificity values. Excess results were present across all metrics at or just after multiples of five and ten. Clear examples of thresholding were seen in the sensitivity value at 50, 60, 70, 75 and 80 (Figure 3). While the specificity had larger excess results at 80 and 90 (Figure 4).

**Figure 1.**
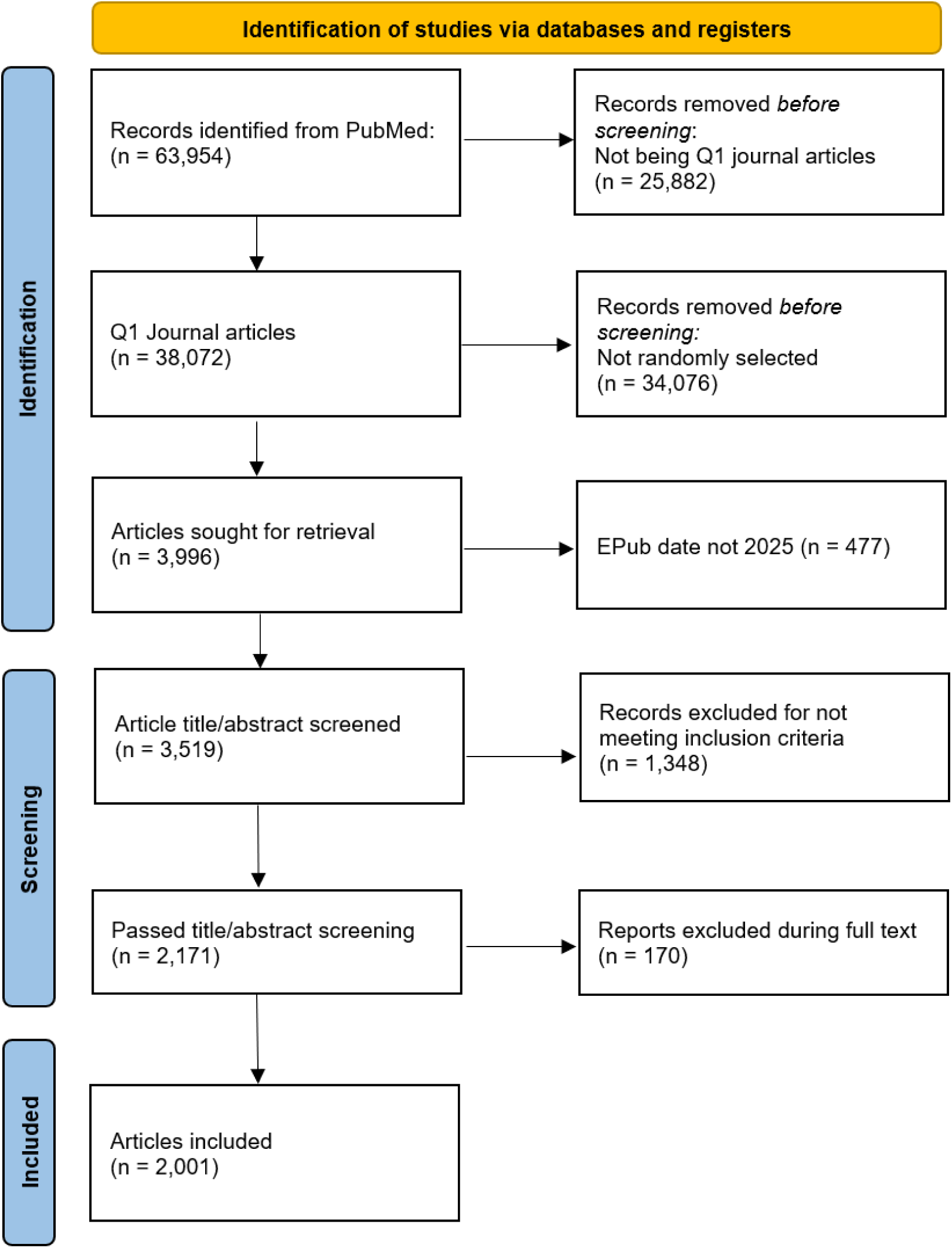
Flow chart of the identification and screening of all included articles in the study.

**Figure 2.**
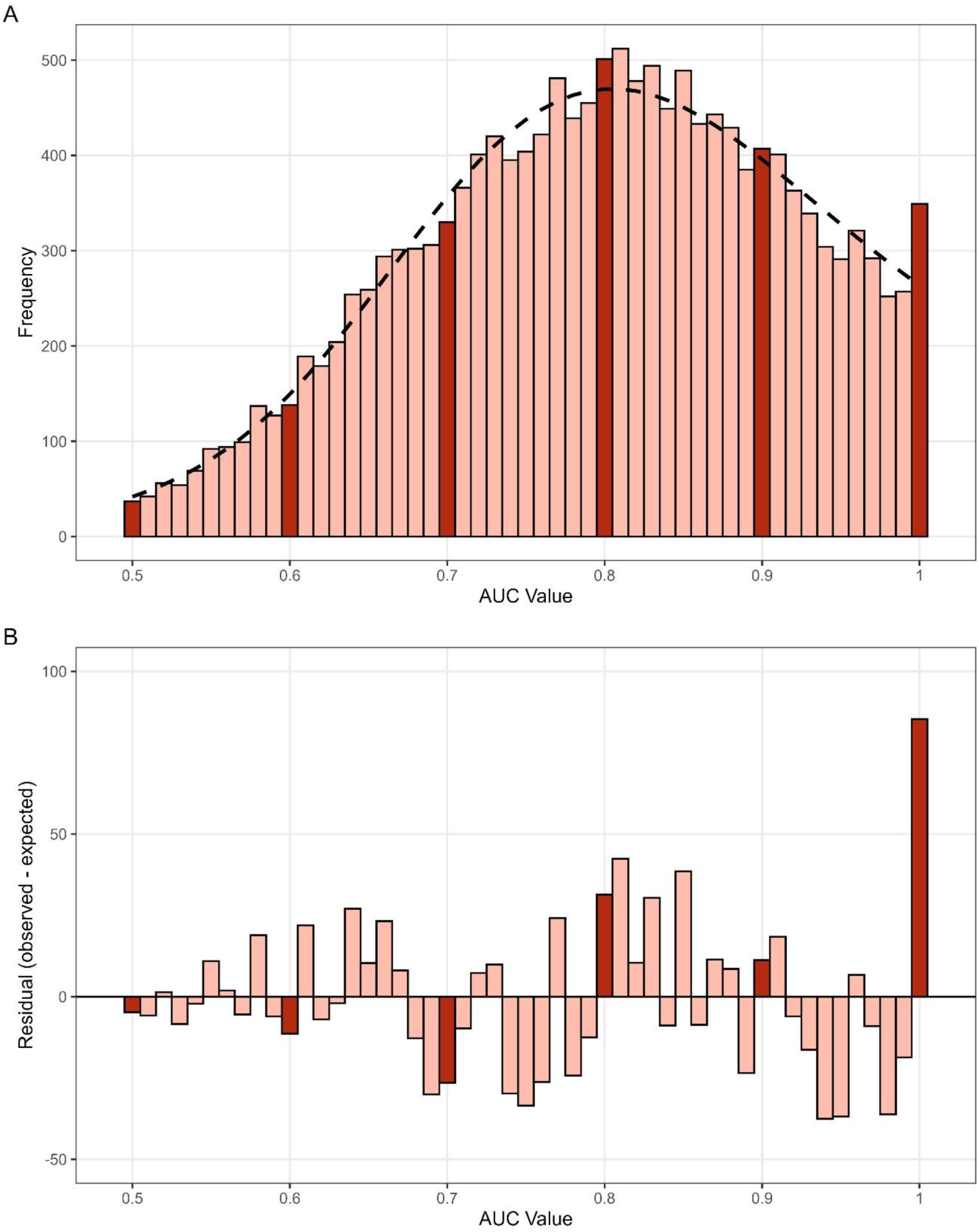
Distribution of all AUC values and the best fitting smooth distribution (dotted line; Panel A). For example, 0.6 on the histogram contains values (0.59, 0.60]. The residuals calculated from the smooth distribution (Panel B). The darker red bars are at multiples of 0.1 which are likely performance thresholds.

**Figure 3.**
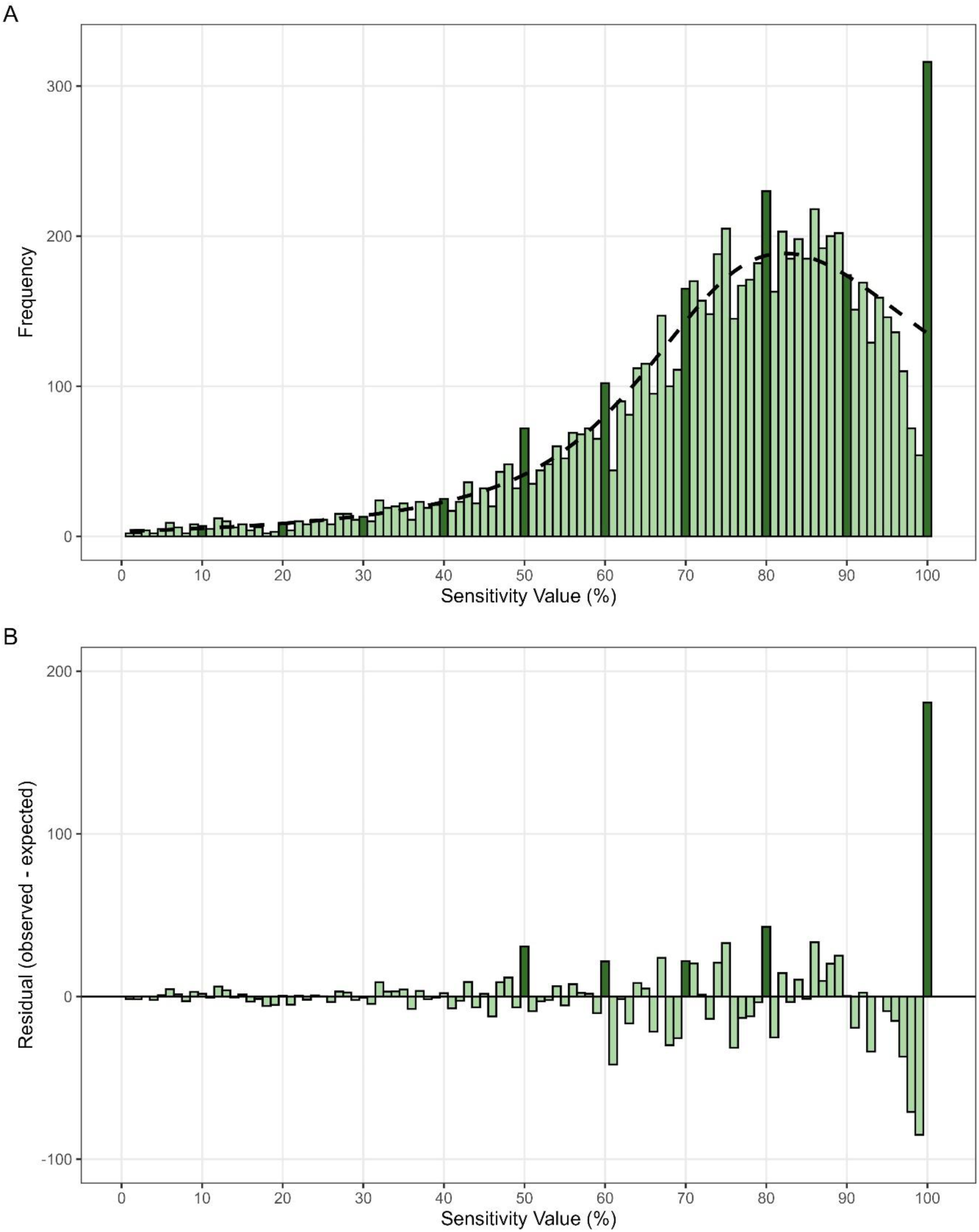
Distribution of the sensitivity values in bin widths of 1% and the best fitting smooth distribution (dotted line; panel A). For example, 60% on the histogram contains values (59%, 60%]. The residuals using the smooth distribution (panel B). The darker green bars are at multiples of 10% which are likely performance thresholds.

**Figure 4.**
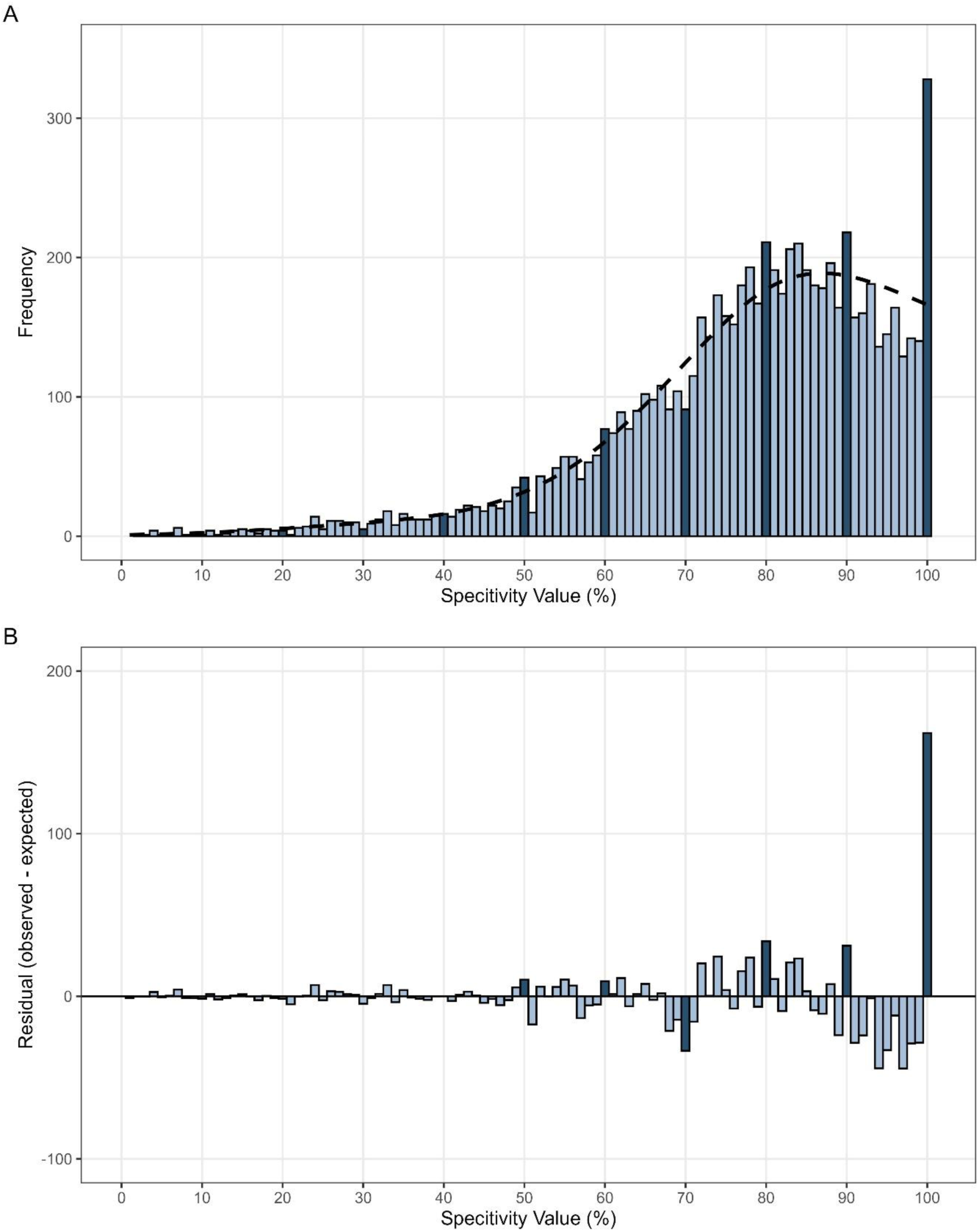
Distribution of the specificity values and the best fitting smooth distribution (dotted line; panel A). The residuals from the best fitting smooth distribution (panel B). The darker blue bars are at multiples of 10% which are likely performance thresholds.

We assumed where both the manual and LLM extractions had the same data, it was correctly extracted. Both methods of extraction contained a similar number of missed or incorrect values. We were comfortable with these results and expect that a purely manual extraction would give similar results to our LLM-based results (more validation information is in Supplementary 3).

### 3.2 ROC Curves

From 200 ROC curves, 189 (95%) AUC values were able to be calculated while 11 (5%) of the ROC curves overlapped with other curves or the figure quality was too poor for accurate extraction. There was no evidence that researchers were systematically altering the reported AUC value compared to the ROC curve data with a mean (SD) difference between the AUC and ROC of 0.0017 (SD = 0.029) (Figure 5). Twelve AUC values differed by less than −0.02 and 14 AUC values differed by more than 0.02 or roughly 1 in 8 ROC curves differed between the AUC value reported in-text and visualised in the ROC curve. Extreme differences in reported AUC values and derived AUC values were seen with the biggest differences of +0.19 and −0.15 which are likely to be clinically important differences.

**Figure 5.**
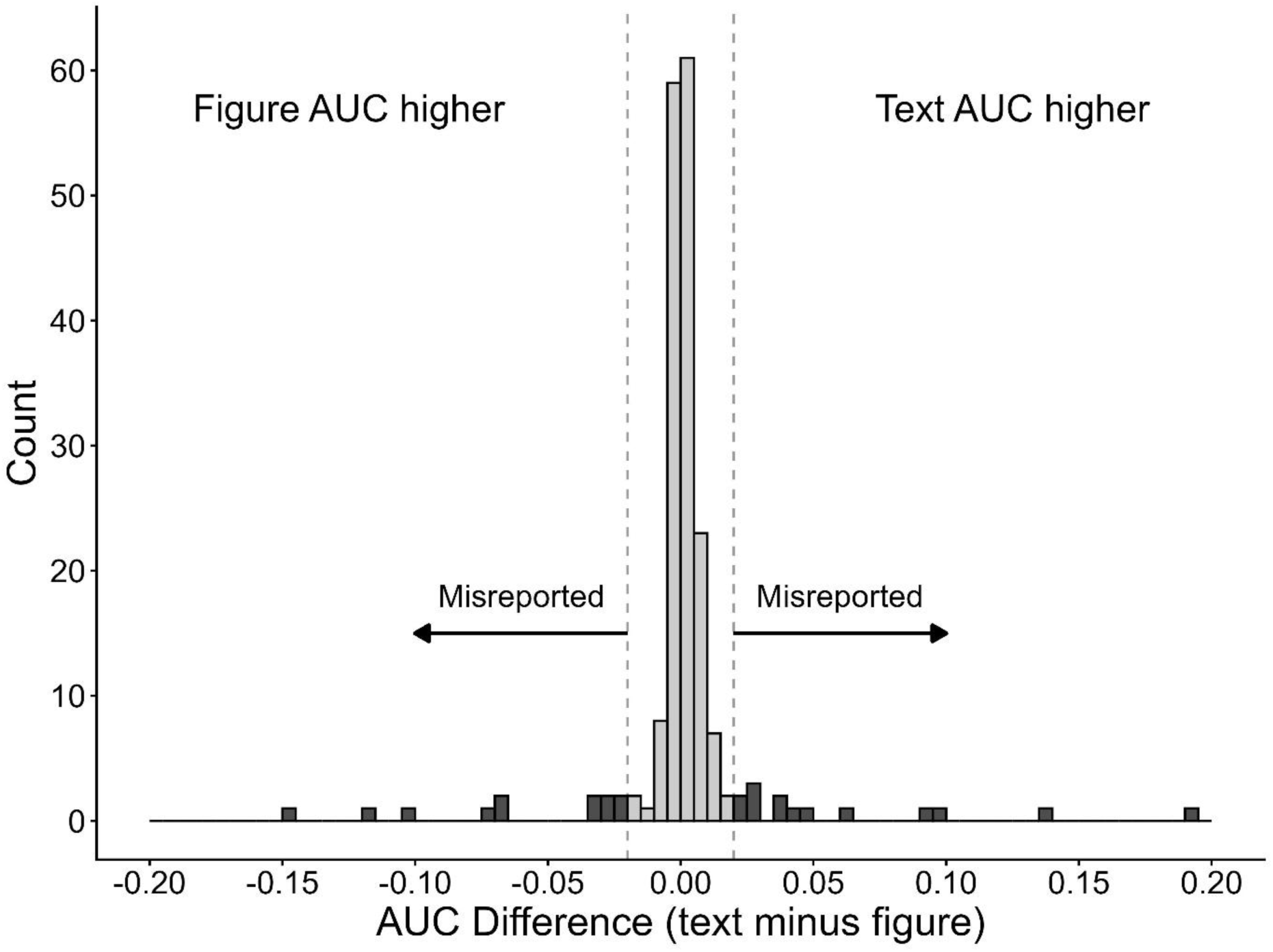
The distribution of differences between the reported AUC value and AUC value from the corresponding ROC curve from a random sample of 187 ROC curves. Bin widths of 0.005. Dark grey bars indicate values that are misreported from the ROC curves while light grey bars indicate values that are reported within the 0.02 threshold.

## 4.0 Discussion

Our results support previous research that performative AUC values are likely being targeted in published clinical prediction model research identified by excess results at arbitrary thresholds. We also found that the sensitivity and specificity are being targeted based on observed excesses at the same thresholds. We are concerned that metrics are being prioritised by researchers for publication at the cost of accuracy. Some of these results could be explained by researchers removing outliers or hard to predict patient subgroups, selectively reporting models that overfit the data. The excess results could also partially be explained by publication bias if these models at purported thresholds are preferentially published. However, we find this less suggestive from the data as the sensitivity and specificity additionally show evidence of being targeted and these excess results all occur across multiple thresholds. This is further supported by a symmetric distribution of reporting errors for AUC values against their corresponding ROC curves (Figure 5), and no evidence was present that researchers are actively increasing AUC values in-text.

These excess results combined with an underrepresentation of expected values between 0.90 and 0.99 show these models close to perfect prediction may have been hacked towards 1.00. Small sample sizes are a common problem and could cause models to overfit and may explain some of these excess models (37–40). The large excess results at (99,100] for the sensitivity and specificity could, however, be explained by rule-in or rule-out models that are designed to be highly selective for sensitivity or specificity but not the other. We find the results near 1.0 for AUC values concerning as these higher performing models may be the ones that make their way to practice as they are perceived to be the most accurate and provide a misleading evidence base for clinicians. These models may be contributing to a medical mirage, appearing numerically excellent on paper but performing less well in practice and misguiding clinical decisions if implemented.

We found that poor reporting practices still persist, despite more than a decade of the TRIPOD+AI reporting guidelines for clinical prediction model research (13). From our analyses we are unable to determine why incorrect reporting continues, but we speculate researchers are developing numerous models and misreporting them potentially via poor version control. The median number of AUC values per article was 5 (1^st^ and 3^rd^ quartile = 3 to 10) which would support this idea of many models. Poor reporting practices may also be diluting the literature of models that have real abilities to improve care and decisions. This situation may reduce the visibility of genuine, high-quality models that may not perform as well but are still clinically useful for their intended use setting (41)

Accurate reporting and representation of clinical prediction models is critical for clinicians to understand expected patient outcomes. We found one model with an AUC value from the ROC curve that was 0.19 worse than the in-text reported value, and another model with an AUC value 0.15 better than the in-text reported value. Potentially serious and substantial consequences could occur if a model with incorrectly reported performance were used to inform policy or care. As 1 in 8 models differed in their underlying data and reporting, and model performances reduce when validated or implemented, there is a serious misrepresentation of research occurring in prediction models.

Clinical prediction models are used in practice, influence clinical decisions and thus patient care and outcomes (42,43). However, clinical guidelines rarely have guidance for appraisal and implementation of prediction models (44), making it plausible for adverse outcomes if these hacked models make their way to practice. Overall, our results indicate 1 in 8 models have AUC values ≥ 0.02 or ≤ 0.02 than in-text reported performances. Differences between reported and underlying performances will misguide clinicians and, if acted upon, potentially lead to harming patients. Combined with excess results near perfect prediction and knowing developmental models typically perform better on paper than in validation studies, results in practice could be substantially worse. We are concerned that prediction models will be making their way into practice which may contribute to worse outcomes for patients. Journals should not recommend but mandate future prediction model development, validation and impact assessments be reported in full against the TRIPOD+AI guidelines unless justified otherwise. While policy makers should prioritise real world evidence from patient outcome assessments to direct the use of prediction models in practice (44).

### 4.1 Limitations

Both the manual extraction and LLM extraction made errors and missed valid data. As with previous research on LLM use for data extraction, many of these errors by both extraction methods arose from poor reporting of results (32). For example, the AUC value was able to be collected with few errors for both manual and LLM extraction, yet large errors were seen in both manual and LLM extraction for sample size. Sample size errors were seen when reported as multiple sub-samples of the entire sample, differing splits for development and validation, or many subsamples during inclusion and exclusion decisions. This extraction failure is not surprising, as clinical prediction models are poorly reported (13). For data extraction to be improved, standardised reporting guidelines for machine readable articles could be used.

We acknowledge that since our sample is based on Q1 journals, some the excess results may be due to publication bias and lower performing models were either not submitted to journals or never passed peer review (45). However, Q1 journals disproportionately publish prediction research and while publication bias would be a component of inflated values this is unlikely to explain the extent of inflated values at the thresholds.

Only including research from 2025 limits our results in time. Information across years may be important to understand changes of hacking practices. Future research examining changes in performance reporting over time could identify if these excess results at thresholds change with the growing use of AI to analyse data and write papers (46).

## 5.0 Conclusions

We found an excess of models with perfect performance values which indicates questionable practices, possibly motivated by a need to oversell model performance. We further suspect these hacking practices are not due to alteration of in-text reporting, rather, hacking practices occur during the analysis based on our examination of ROC curves. We expect these hacking practices primarily to come from data or methodological manipulations that inflate model performances over publication bias. One in eight models incorrectly report the AUC value compared to the ROC curve, with some substantially and potentially clinically imported differences. Broadly, we identify that performance metrics in clinical prediction models are being targeted and highlight poor reporting is extensive in clinical prediction model research with more than two-thirds of articles not reporting confidence intervals and 1 in 40 AUC values with confidence intervals are errors. Our study suggests that paper-based performance metrics may be prioritised over practical care, and this could be harming patients. Alternatively, if none of these models made their way into practice, it may constitute an enormous volume of research waste.

## Data Availability

All data produced are available online at GitHub (https://github.com/alexdgibson/cpm_auc).

