## Supplementary 1 for "Clinical Prediction Model Performance Metrics Cluster at Arbitrary Thresholds and are Incorrectly Reported in High-Ranking Journals: An Observational Meta-Research Study"

Search filter: "((((((((((((((((c-stat[Title/Abstract]) OR (c-statistic[Title/Abstract])) OR (c-index[Title/Abstract])) OR (concordance stat[Title/Abstract])) OR (concordance statistic[Title/Abstract])) OR (auc[Title/Abstract])) OR (auc value[Title/Abstract])) OR (area under the curve[Title/Abstract])) OR (area under the curve value[Title/Abstract])) OR (area under the receiver operating characteristic curve[Title/Abstract])) OR (receiver operating characteristic curve[Title/Abstract])) OR (area under the receiver operating characteristic curve value[Title/Abstract])) OR (receiver operating characteristic curve value[Title/Abstract])) OR (roc-auc[Title/Abstract])) OR (roc curve[Title/Abstract])) OR (roc curve value[Title/Abstract])) NOT (pharmacokinetic[Title/Abstract])"
