## Supplementary 2 for "Clinical Prediction Model Performance Metrics Cluster at Arbitrary Thresholds and are Incorrectly Reported in High-Ranking Journals: An Observational Meta-Research Study"

Prompt used for data extraction in Claude Sonnet 4.6:

You are conducting data extraction for a medical research study. Your task is to accurately extract all data from PDF clinical prediction model articles and output a CSV file.

Step 1 — Identify all models first. Before extracting any data, read the entire article and list every clinical prediction model reported. Models can differ by predictors, method/algorithm, outcome, or dataset (e.g. development vs validation cohort), collect all models even if they differ in these variables. There could be one model in an article or tens of models. Carefully find all models. There can be articles which include development, and validation models which can differ in sample sizes. Data can be in-text, in tables or in figures, use all this information. Number them in the order they appear in the text. Where the same model is applied to different outcomes or methods (e.g. 15-year, 20-year, 25-year mortality or logistic regression, k-nearest neighbours, etc), treat each model combination as a separate model.

Step 2 — Extract data for every model. For each model identified in Step 1, extract the following variables. Output only the values — no commentary. CSV file columns (output exactly this header row): doi,article,model,sample_size,study_type,model_type,auc,auc_ci_low,auc_ci_upper,sens,sens_ci_lower,sens_ci_upper,spec,spec_ci_lower,spec_ci_upper,roc,roc_colour,roc_panel.

Variable definitions:

- doi — this is the PDF filename only use the pdf file name, as a character variable. Do not change this.
- article — article number in the list of articles being processed as a numeric variable
- model — model number within the article, ordered as reported in text as a numeric variable
- sample_size — sample size for the specific model (may differ across models in the same article) as a numeric variable
- study_type — one of: development, internal validation, or external validation. As a character variable
- model_type — diagnostic or prognostic as a variable as a character variable
- auc — area under the curve, AUC or AUROC value for this model as a variable as a numeric variable
- auc_ci_low — lower bound of the AUC confidence interval as a numeric variable
- auc_ci_upper — upper bound of the AUC confidence interval as a numeric variable
- sens — sensitivity for the model (must correspond to the same threshold/cutpoint as spec) as a numeric variable
- sens_ci_lower — lower bound of sensitivity confidence interval as a numeric variable
- sens_ci_upper — upper bound of sensitivity confidence interval as a numeric variable
- spec — specificity for the model (must correspond to the same threshold/cutpoint as sens) as a numeric variable
- spec_ci_lower — lower bound of specificity confidence interval as a numeric variable
- spec_ci_upper — upper bound of specificity confidence interval as a numeric variable
- roc — yes or no: was an ROC curve figure reported for the model's AUC value? as a character variable
- roc_colour — colour of the ROC curve corresponding to the model's AUC value as a character variable
- roc_panel — figure number and panel label (e.g. Figure 2A) where this model's ROC curve appears as a character variable
- exclude — yes if the article is not a clinical prediction model study, otherwise no as a character variable

Rules:

- Extract data for all models identified and only extract the data specified.
- All values for a given row must belong to the same model.
- If a value is not reported in the article, enter NA.
- If it cannot be determined which ROC curve corresponds to which AUC value, enter "could not determine" for roc_colour and roc_panel.
- Do not skip any model. Do not add explanatory text outside the CSV.
- Output begins with the header row, followed by one row per model.
- Ensure the final output is a .csv file
