## Supplementary 3 for "Clinical Prediction Model Performance Metrics Cluster at Arbitrary Thresholds and are Incorrectly Reported in High-Ranking Journals: An Observational Meta-Research Study"

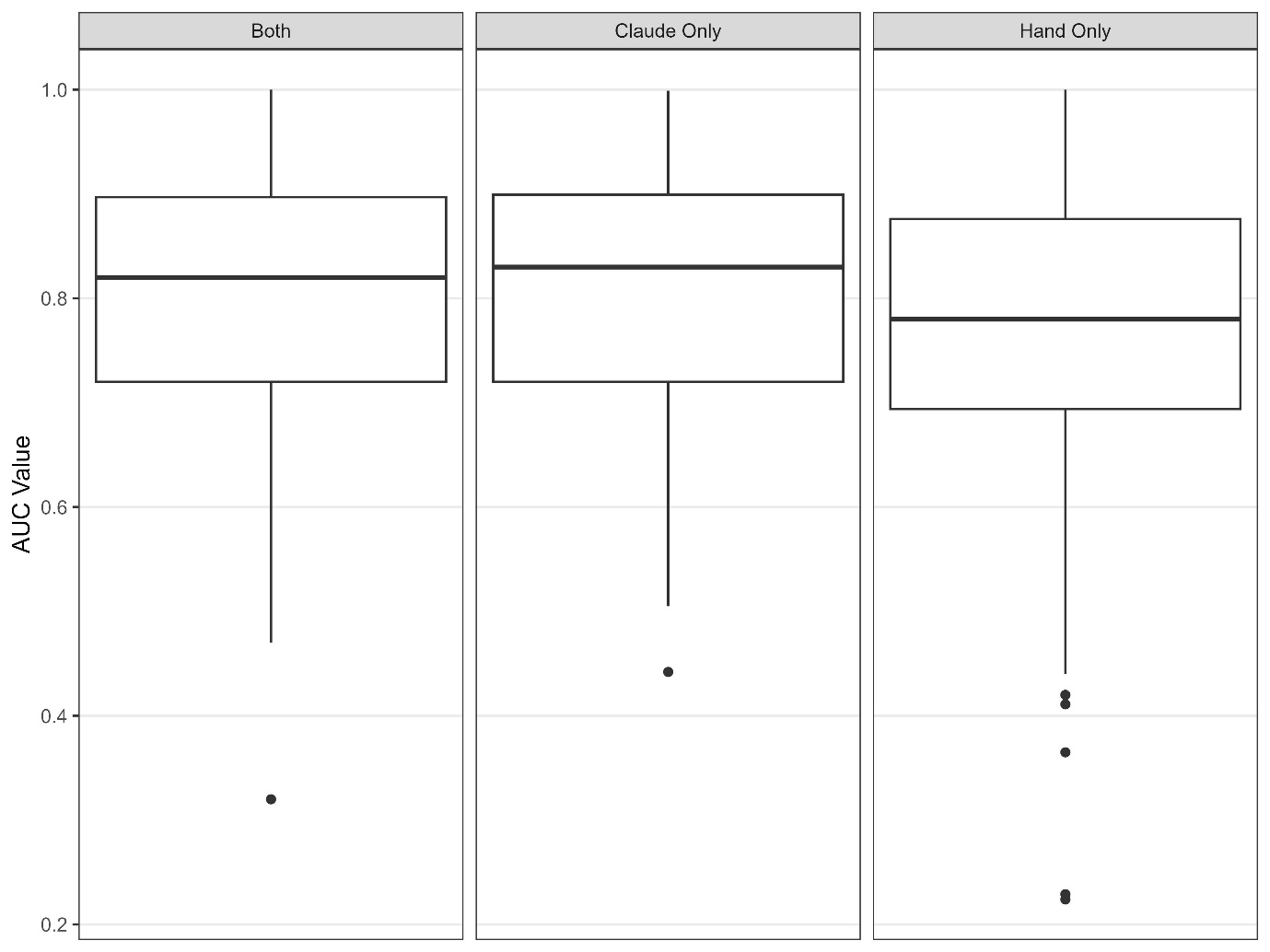

Figure 2. Box plot for AUC values where both the LLM and manual extracted the same values, where the LLM only collected values, and where manual only collected values. Both contain 1,045 AUC values, LLM only contains 310 AUC values and manual only contains 485 AUC values.

As there is no gold standard for data extraction we validated where errors may be present. We took 50 randomly selected values where only the manual extraction retrieved data, and the LLM extraction did not, and vice versa (Table 1). Validation showed the LLM extraction had comparable performance to manual extraction (Table 1).

|  | AUC | Sensitivity | Specificity |
| --- | --- | --- | --- |
| Manual and LLM retrieved | 1,045 | 397 | 395 |
| Manual only retrieved | 485 | 185 | 187 |
| LLM only retrieved | 310 | 254 | 216 |
| Validation Checks (number correct/50) |  |  |  |
| Manual only | 48/50 | 48/50 | 47/50 |
| LLM only | 50/50 | 49/50 | 47/50 |

**Table 1.** Comparison and validation for manual data extraction and LLM data extraction from 176 articles of 1,325 LLM extracted articles. For example, manual extraction retrieved 485 AUC values the LLM did not, and the LLM retrieved 310 AUC values manual extraction did not. AUC values extracted values the manual extraction had 2 errors, and the LLM extraction had 0 errors.

| Degrees of Freedom (edge knots) | AUC | Sensitivity | Specificity |
| --- | --- | --- | --- |
| 2 | 85470.1 | 49506.3 | 45856.8 |
| 3 | 85415.3 | 49446.6 | 45730.5 |
| 4 | 85415.3 | 49365.7 | 45696.4 |
| 5 | NA | 49370.6 | 45694.9 |
| 6 | NA | 49364.4 | 45691.3 |
| Degrees of Freedom (distance knots) |  |  |  |
| 2 | 85429.7 | 49460.4 | 45769.5 |
| 3 | 85415.3 | 49461.3 | 45751.4 |
| 4 | 85415.3 | 49361.9 | 45702.8 |
| 5 | NA | 49362.7 | 45689.5 |
| 6 | NA | 49364.4 | 45691.3 |

**Table 2.** The Akaike information criterion (AIC) to assess best fit for the smooth distribution. All values are rounded to one decimal place. The best fitting smooth used three degrees of freedom for the AUC with limited knots, four degrees of freedom for the sensitivity and five degrees of freedom for the specificity with knots outside the 0 to 100% limits.
